# Factors related to the performance of the periodontal specialty in secondary oral health in Brazil

**DOI:** 10.1101/2023.06.06.23291027

**Authors:** Vinícius de Moraes Simião, Estéfany Figueiredo Gonzalez, Livia Fernandes Probst, Rafaela da Silveira Pinto, Alessandro Diogo De-Carli

**Affiliations:** Postgraduate Program in Family Health, Universidade Federal de Mato Grosso do Sul, Campo Grande, Mato Grosso do Sul, Brazil; Multiprofessional Residency Program in Family Health SESAU/Fiocruz, Campo Grande, Mato Grosso do Sul, Brazil; Health Technology Assessment Unit. Hospital Alemão Oswaldo Cruz, São Paulo, São Paulo, Brazil; Department of Social and Preventive Dentistry, Universidade Federal de Minas Gerais, Belo Horizonte, Minas Gerais, Brazil; Faculty of Dentistry, Universidade Federal de Mato Grosso do Sul, Campo Grande. Mato Grosso do Sul, Brazil

**Keywords:** Secondary Care, Periodontics, Dental Specialty, Dental Health Services, Health Services Research

## Abstract

**Objective:** The aim of this study was to investigate, at a national level, which individual factors of the work process/infrastructure are associated with the achievement of goals in the periodontics specialty in Brazilian Dental Specialty Centers (CEO).

**Methods:** This was a quantitative, analytical, cross-sectional study. Secondary data from DATASUS and the external evaluation of the second cycle of the CEO Access and Quality Improvement Program were used. Variable description was carried out in the first stage, and then the bivariate Poisson regression was performed to verify possible associations between the variables and the outcome (achievement of goals in Periodontics in the CEO). In this analysis, the covariates that were associated with the outcome at the p <0.20 significance level were included in the next step of the analysis. Multivariate Poisson regression with a robust estimator was then performed with those that met the above criterion. The variables that showed a p value < 0.05 were considered in the final model.

**Results:** The outcome was achieved in more than seven months of the year (mean 7.03 months, SD 4.20). Most CEO monitored the established goals (93.2%), had referral as the only way of access (61.7%), had only municipal coverage (68.4%), carried out planning and periodic evaluation of actions (89.2%). A minority has quotas of procedures by Oral Health teams (OHTs) in Primary Health Care (PHC) (18.8%). The presence of a specialist in periodontics was (on average) 1.16 per CEO and the sum of the workload of dentists working in this specialty was 31.1 hours (SD = 23.9).

**Conclusion:** It was concluded that the individual factors of the work process/infrastructure associated with the achievement of goals in periodontics in Brazilian CEO are: monitoring of established goals, CEO scope and number of professionals working in the specialty.

## Introduction

With the implementation of the National Oral Health Policy (PNSB, *Política Nacional de Saúde Bucal*) in 2004, by the Ministry of Health (MoH), oral health care in the Brazilian Unified Health System (SUS, *Sistema Único de Saúde*), historically characterized by difficult access and limited to mutilating techniques, started to include the promotion, prevention and recovery of oral health of the Brazilian population. Therefore, the reorganization and qualification of the service provided by the SUS was crucial (1,2).

From this perspective, the PNSB promoted the expansion of the coverage provided by the oral health teams (OHTs) in the Family Health Strategy (FHS) to reorganize access to oral health in Primary Health Care (PHC) and implemented the Dental Specialty Centers (CEO, *Centros de Especialidades Odontológicas*), aiming to expand access to Secondary Oral Health Care (SOHC) (1,2). The CEO are health care establishments listed in the National Register of Health Establishments (CNES, *Cadastro Nacional de Estabelecimentos de Saúde*), which are obliged to offer basic oral health services in the Oral Diagnosis, Advanced Periodontics, Minor Oral Surgery, Endodontics and Dental Care specialties to patients with special needs. The granting of funding for the CEO maintenance is linked to the achievement of goals by the specialty (1).

The CEO are classified according to their composition regarding the number of dental chairs in the establishment. Therefore, Modality I, II and III of the CEO consist of three, four to six, and more than seven dental chairs, respectively. These establishments operate for 40 hours a week and the number of professionals working there varies according to the modality, with the achievement of goals being monitored by the MoH (3).

However, in addition to offering greater access to SOHC, it was necessary to invest in evaluation processes at this level of care, aiming to attain better quality of the services offered by the CEO. For this purpose, the MoH implemented the CEO component in the National Access and Quality Improvement Program (PMAQ, *Programa Nacional de Melhoria do Acesso e da Qualidade*) through Ordinance GM/MoH N. 261, of February 21, 2013, having its rules revised by Ordinance GM/MoH N. 1,599, of September 30, 2015 (3). The PMAQ-CEO, until 2018, corresponded to the federal initiative for the evaluation and monitoring of the SOHC in Brazil.

This study aims to assess the achievement of the goals in the periodontics specialty of Brazilian CEO. In this specialty, the goal to be achieved varies according to the type of CEO, comprising 60 procedures for CEO type I, 90 for type II and 150 for type III (4), which were verified by the PMAQ-CEO evaluation process in its second cycle, in 2018 (5)

The epidemiological picture of periodontal conditions in Brazil is still considered inconclusive (6). However, if one considers the CDC/AAP (Centers for Disease Control/ American Academy of Periodontology) criteria, the prevalence of periodontal disease in Brazil is still higher than in developed countries (6). In the United States, the prevalence of severe periodontitis ranges from 6.7 to 11.7%, while in Brazil it ranges from 34.4% to 63.8% in individuals aged 35 years or older (6).

The need to analyze compliance with periodontics goals in the CEO is justified, as there are no studies in this area with this specific approach, making it an unprecedented analysis. Moreover, it is necessary to consider the critical role of CEO in relation to the attention/control of periodontal disease, considering that, in terms of public policy, it corresponds to the only access for most Brazilians to periodontal care at a specialized level, via SUS.

The aim of this study is to investigate, at a national level, which individual factors are associated with the achievement of goals in the periodontics specialty in Brazilian CEO.

## Method

### Ethical aspects

The microdata used in this study were obtained from National Information Systems with public and unrestricted access.

The PMAQ-CEO was approved by the Research Ethics Committee (CEP) of the Federal University of Pernambuco (UFPE), under CAAE number: 23458213.0.0000.5208, complying with the requirements of Resolution n. 466/12 of the National Health Council.

### Study design and context

This is an analytical cross-sectional study using secondary data and reported in accordance with the Strengthening the Reporting of Observational Studies in Epidemiology (STROBE) statement (7). The study uses data from the external evaluation of the 2^nd^ cycle of the National Program to Improve Access and Quality of Dental Specialty Centers (PMAQ-CEO), carried out in 2018, in Brazil (3). Microdata related to modules I and II of the PMAQ-CEO were used, which evaluated the structure and work process in these establishments, respectively.

### Study universe and sample

The universe of the study comprises the municipalities in which the CEO adhered to the PMAQ and who answered the external evaluation questionnaire. Of the CEO implemented in the Brazilian territory in 2018, 1097 answered the external evaluation questionnaire. Of these, 104 were excluded for showing zero production in all months of the year, 15 for not having identified the type of CEO and 05 for not providing individual data in the database of the PMAQ-CEO 2^nd^ cycle. Therefore, the final sample consisted of 973 CEO from 809 municipalities in Brazil. The authors did not had access to information that could identify individual participants during or after data collection

### Analyzed variables

The variables used in the study are described in Chart 1.

**Chart 1.** Variables (outcome and independent) used in the study.

| Variable | Categories | Data source |
| --- | --- | --- |
| **Outcome** |  |  |
| Number of months in which the goal was achieved in the periodontics specialty | Achieved the goal.<br>Did not achieve the goal. | DATA-SUS |
| **Independent variables of the 1<sup>st</sup> level: related to CEO.** |  |  |
| Type of access | Referral only.<br>Mixed (spontaneous and referral) | PMAQ-CEO |
| Quotas of procedures per oral health team in PHC | Yes<br>No | PMAQ-CEO |
| Monitoring of established goals | Yes<br>No | PMAQ-CEO |
| Sum of periodontics workload | Description (Number) | PMAQ-CEO |
| Number of dentists working in periodontics | Description (Number) | PMAQ-CEO |
| Planning and periodic evaluation of actions | Yes<br>No | PMAQ-CEO |
| CEO scope | Yes<br>No | PMAQ-CEO |
| CEO type | Type I<br>Type II<br>Type III | PMAQ-CEO |
| Legend: CEO = Dental Specialty Centers; PHC = Primary Health Care; PMAQ-CEO = National Access and Quality Improvement Program of Dental Specialty Centers |  |  |

### Data organization and statistical analysis

All secondary data were organized in spreadsheets that comprised the researcher’s own database, using the Microsoft Excel program. All statistical analyses were performed using the SPSS software, version 23.0. Variable description was performed in the first stage, and then the bivariate Poisson regression analysis was used to verify possible associations between the independent variables and the outcome. In this analysis, the covariates that were associated with the outcome at the significance level of p-value <0.20 were included in the next step of the analysis. Poisson multivariate regression with a robust estimator was then performed with those variables that met the previous criterion. The variables that reached a p-value <0.05 were considered in the final model.

## Results

Overall, the CEO reached goals in the periodontics specialty in more than seven months of the year (mean: 7.03 months, SD: 4.20). Most CEO monitored the established goals (93.2%), had referral as the only way of access (61.7%), had municipal coverage only (68.4%), and carried out planning and periodic evaluation of actions (89.2%). A minority has quotas for OHT procedures in the PHC (18.8%). The presence of a specialist in periodontics was (on average) 1.16 per CEO and the sum of the workload of dentists working in this specialty was 31.1 hours (SD = 23.9) (Table 1).

**Table 1.**
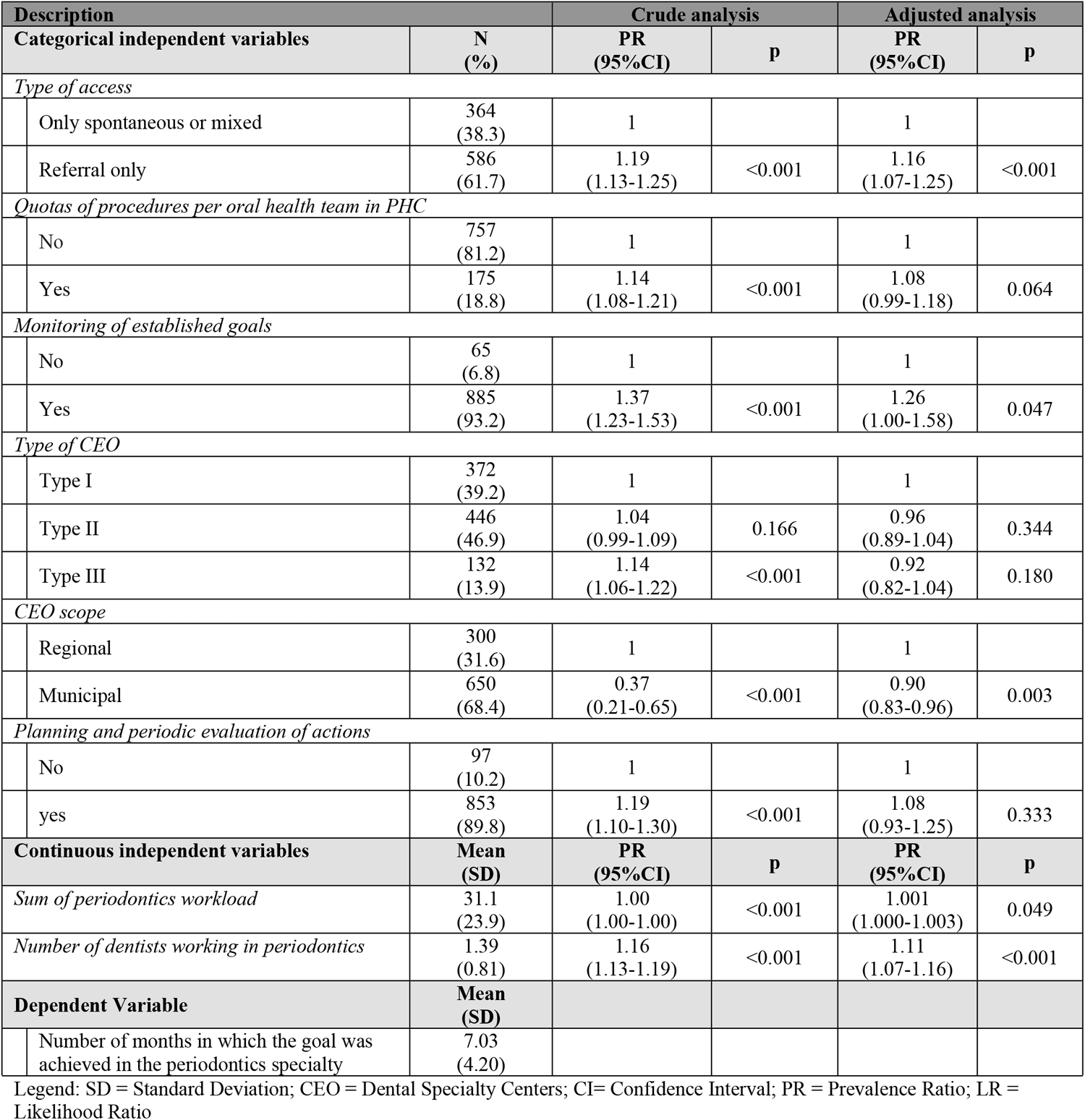
Analysis of independent variables and study outcome.

Table 1 shows the analysis for the outcome (number of months in the year in which the CEO reached their goals in the periodontics specialty). Poisson regression showed this outcome was associated with variables related to the individual level of dental specialty centers. The CEO that monitors their established goals were 1.37 times more likely to achieve these goals than the ones who did not monitor the established goals. In turn, a higher number of dentists who work in periodontics and CEO with regional coverage were also variables associated with the achievement of goals.

## Discussion

The findings of the present study demonstrate an association between the achievement of goals in the specialty of Periodontics and factors that are proximal to the work process in Brazilian CEO.

To date, no publications have been identified with the same research object. Therefore, the information disclosed here and the problematization of the results contribute to the advancement of the construction of knowledge in the specific field of SOHC assessment in Brazil, having Periodontics as a guiding axis.

Different individual factors were identified, which are related to the number of months in which goals were met in Periodontics in Brazilian CEO. Therefore, the monitoring of the established goals, the CEO scope and the number of dentists working in the specialty suggest that issues intrinsic to the work process can contribute to a better performance of these establishments.

The monitoring of goals was significantly associated with the tendency to achieve them, since the monitoring and planning of the provided services are of the utmost importance to achieve good results in the quality indicators and standards (3). In terms of evaluation, this becomes relevant, considering that procedural issues inherent to the daily routine of the services can be used as support to guide the CEO team regarding the process of negotiating and contracting goals with managers, as well as defining priorities for improving the service quality based on the recognition of the achieved results, whether they are effective or in need of improvement regarding the intervention strategies (3).

As the establishments that are regional referrals were more successful in meeting the goals, it is plausible that, in these CEO, as they are located in referral municipalities in the health regions, there may be better infrastructure, organization of patient flow and demand, as well as of the Oral Health Care Network. These factors may contribute to better results in relation to the outcome, when compared to CEO of municipal scope.

With regard to human resources, the number of dentists who work in the specialty of periodontics in CEO showed to be an important factor for the achievement of goals. The results show that, on average, there is more than one periodontist per CEO, which may be an assumption for the high number of procedures performed in this specialty (8).

The data presented herein are relevant because, in addition to being unprecedented, they can be used as guidelines for the planning and implementation of specific actions in the field of Periodontics, considering that a large part of the Brazilian population depends on public health services, especially in secondary oral health care (9). In this context, this becomes even more important, as it is known that data related to the prevalence of periodontal disease in Brazil are unequal (10). This can be justified by the characteristics of the country, such as diversity in socioeconomic, demographic, environmental and behavioral factors originating from its large territorial extension (11), which can be impacted by the lack of a care protocol and standardized criteria for characterizing the disease (12) in the SUS.

Regardless of the discrepancy of results, in a study carried out using the Centers for Disease Control/American Academy of Periodontology (CDC/AAP) criteria, considering the limit of clinical attachment loss ≥ 3mm, it is suggested that Brazil has a higher prevalence of major periodontal diseases than that found in more developed countries, ranging from 34.4% to 63.8% in individuals aged 35 or over (6).

Thus, it indicates a challenge for the PNSB in minimizing limitations, especially those related to the work process, so that access to quality and effective services in periodontics can be promoted, impacting the oral health of the population. In this sense, it is necessary to resume the planning of implementation actions aimed at individuals/communities in a situation of greater risk and vulnerability, which is already foreseen in the PNSB (1), but which, since 2016, with the implementation of the fiscal austerity policy, has been suffering cutbacks and devaluation at a national level.

This has become a critical issue, as it is known that, in a country as unequal as Brazil, there is still an association between socioeconomic variables and the achievement of goals in periodontics, where the size of the population and the Municipal Human Development Index (MHDI) are associated with the CEO performance (13,14), and there is a correlation between contextual and socioeconomic factors, suggesting inequality in the need for periodontal treatment in the Brazilian elderly population (15).

In this study, the number of months in which the goal of procedures was reached in the specialty of periodontics was, on average, more than seven months. The growing trend of CEO that meet periodontics goals may be related to the investment in oral health actions from 2003 to 2014, when the PNSB provided conditions to expand the supply and the capacity of services provided by this specialty (16).

This study has some limitations. Despite the representativeness of the sample, these data should be generalized with caution, as the CEO participation in the PMAQ-CEO evaluation process was not mandatory, but voluntary. Secondary data obtained from national information systems with public and unrestricted access offer numerous advantages, such as broad population coverage and low cost for information collection. However, as these data are usually collected in routine health services without a *priori* research purposes, the absence of important information for the analyses of interest can represent significant disadvantages (17).

In turn, the quality of information from databases can be assessed in two dimensions: completeness and accuracy. Completeness refers to the extent to which data are missing from the perspective of the outlined research question. Missing data is unavoidable; however, it is often necessary to understand the extent to which important variables are missing and the possible reasons for their absence. Another important dimension is accuracy. Information from electronic system records, such as procedure codes or numeric values, can sometimes be recorded inaccurately (17).

From an investigative perspective, it is suggested that new studies be carried out, exploring the interface between the Periodontics actions implemented in the PNSB, PHC and the use of the service by the user. Therefore, studies using mixed methods are essential, which have the power to go beyond quantitative generalizations, further analyzing critical issues inherent to the complexity of the health-disease process in the context of the SUS.

## Conclusion

It was concluded that the factors associated with the achievement of goals in periodontics in Brazilian CEO include the monitoring of established goals, the CEO scope and the number of dentists working in the specialty.

## Data Availability

The study uses data from the external evaluation of the 2nd cycle of the National Program to Improve Access and Quality of Dental Specialty Centers (PMAQ-CEO), carried out in 2018, in Brazil. Available from: https://189.28.128.100/dab/docs/portaldab/documentos/microdados_pmaq/documentos/Banco_2CICLO_PMAQ-CEO.zip

## References

1. BRASIL. Ministério da Saúde. Diretrizes da Política Nacional da Saúde Bucal. Brasília; 2004.

2. Pucca GA, Gabriel M, de Araujo Med, de Almeida Fcs. Ten years of a national oral health policy in Brazil: Innovation, boldness, and numerous challenges. J Dent Res. 2015 Oct 1;94(10):1333–7.

3. BRASIL. Ministério da Saúde. Secretaria de Atenção à Saúde. Departamento de Atenção Básica. Manual instrutivo do Pmaq para as equipes de Atenção Básica (Saúde da Família, Saúde Bucal e Equipes Parametrizadas) e Nasf. Brasília; 2017.

4. BRASIL. Ministério da Saúde. Portaria no. 1464 de 2011. Altera o Anexo da Portaria no 600/GM/MS, de 23 de março de 2006, que institui o financiamento dos Centros de Especialidades Odontológicas (CEO). 2011.

5. Pinto R da S, Lucas SD, Goes PSA de, Silva SL da, Neves ÉSM, Zina LG, et al. Contextual and local determinants associated with the achievement of goals in the endodontics specialty in Brazilian dental speciality centres: A multilevel analysis. Community Dent Oral Epidemiol. 2022 Feb 1;50(1):74–82.

6. Caroline Cunha de Medeiros T, Areas Souza A, Nogueira Haas A, Paulo Steffens J. Epidemiology of periodontitis in Brazilian adults: an integrative literature review of large and representative studies. Vol. 24, Journal of the International Academy of Periodontology 2022. 2022.

7. von Elm E, Altman DG, Egger M, Pocock SJ, Gøtzsche PC, Vandenbroucke JP. The Strengthening the Reporting of Observational Studies in Epidemiology (STROBE) Statement: Guidelines for reporting of observational studies. Notfall und Rettungsmedizin. 2008;11(4):260–5.

8. Pires ALC, Gruendemann JLAL, Figueiredo GS, Conde MCM, Corrêa MB, Chisini LA. Atenção secundária em saúde bucal no Rio Grande do Sul: análise descritiva da produção especializada em municípios com Centros de Especialidades Odontológicas com base no Sistema de Informações Ambulatoriais do Sistema Único de Saúde. Revista da Faculdade de Odontologia - UPF. 2016 May 18;20(3).

9. IBGE. Pesquisa nacional de saúde: 2019: informações sobre domicílios, acesso e utilização dos serviços de saúde: Brasil, grandes regiões e unidades da federação / IBGE, Coordenação de Trabalho e Rendimento.2019.

10. Teixeira FCF, Marín-León L, Gomes EP, Pedrão AMN, Pereira A da C, Francisco PMSB. Perda de inserção periodontal e associações com indicadores de risco sociodemográficos e comportamentais. Rev Odontol UNESP. 2019;48.

11. Silva GCB da, Melo Neto O de M, Nascimento AMV do, Santos CAO dos, Nóbrega Wfs, Souza SLX de. História Natural da Doença Periodontal: uma revisão sistematizada. Research, Society and Development. 2020 Jun 1;9(7):e607974562.

12. Oppermann RV, Haas AN, Rösing CK, Susin C. Epidemiology of periodontal diseases in adults from Latin America. Periodontology 2000. 2015;67(1):13–33. doi:10.1111/prd.12061.

13. Sávio P, de Goes A, Figueiredo N, Cavalcanti Das Neves J, Moura Da F, Silveira M, et al. Avaliação da atenção secundária em saúde bucal: uma investigação nos centros de especialidades do Brasil. Rio de Janeiro; 2012. Available from: http://portal.saude.gov.br/portal/se/da

14. Cortellazzi KL, Balbino EC, Guerra LM, Vazquez F de L, Bulgareli JV, Ambrosano GMB, et al. Variables associated with the performance of Centers for Dental Specialties in Brazil. Revista Brasileira de Epidemiologia. 2014 Oct 1;17(4):978–88.

15. Dalazen CE, de Carli AD, Bomfim RA, Santos MLM dos. Contextual and individual factors influencing periodontal treatment needs by elderly brazilians: A multilevel analysis. PLoS One. 2016 Jun 1;11(6).

16. Andrade FB, Silveira Pinto R, Antunes JLF. Trends in performance indicators and production monitoring in specialized dental clinics in brazil. Cad Saude Publica. 2020;36(9).

17. Liu M, Qi Y, Wang W, Sun X. Toward a better understanding about real-world evidence. European Journal of Hospital Pharmacy. 2022 Jan;29(1):8–11.

